# AutoAudio: Deep Learning for Automatic Audiogram Interpretation

**DOI:** 10.1101/2020.04.30.20086637

**Authors:** Matthew G. Crowson, Jong Wook Lee, Amr Hamour, Rafid Mahmood, Aaron Babier, Vincent Lin, Debara L. Tucci, Timothy C.Y. Chan

**Author notes:** **Corresponding Author:** Matthew G. Crowson MD MPA FRCSC, Sunnybrook Health Sciences Center, 2075 Bayview Avenue, Toronto, Ontario, M4N 3M5, Department of Otolaryngology-Head & Neck Surgery. **First-Author**. **Manuscript Submission:** (1) Each of the authors indicated above have contributed to, read and approved this manuscript. (2) FINANCIAL DISCLOSURE: no authors have disclosures related to this manuscript. (3) CONFLICT DISCLOSURE: no authors have conflicts related to this manuscript. (4) In consideration of the journal reviewing and editing my submission, the authors undersigned transfers, assigns and otherwise conveys all copyright ownership in the event that such work is published.

## Abstract

**Objectives:** Hearing loss is the leading human sensory system loss, and one of the leading causes for years lived with disability with significant effects on quality of life, social isolation, and overall health. Coupled with a forecast of increased hearing loss burden worldwide, national and international health organizations have urgently recommended that access to hearing evaluation be expanded to meet demand.

**Methods:** The objective of this study was to develop ‘AutoAudio’ – a novel deep learning proof-of-concept model that accurately and quickly interprets diagnostic audiograms. Adult audiogram reports representing normal, conductive, mixed and sensorineural morphologies were used to train different neural network architectures. Image augmentation techniques were used to increase the training image set size. Classification accuracy on a separate test set was used to assess model performance.

**Results:** The architecture with the highest out-of-training set accuracy was ResNet-101 at 97.5%. Neural network training time varied between 2 to 7 hours depending on the depth of the neural network architecture. Each neural network architecture produced misclassifications that arose from failures of the model to correctly label the audiogram with the appropriate hearing loss type. The most commonly misclassified hearing loss type were mixed losses.

**Conclusion:** Re-engineering the process of hearing testing with a machine learning innovation may help enhance access to the growing worldwide population that is expected to require audiologist services. Our results suggest that deep learning may be a transformative technology that enables automatic and accurate audiogram interpretation.

## INTRODUCTION

The National Academy of Sciences (NAS) and World Health Organization (WHO) have identified hearing loss as the leading human sensory system loss, and one of the leading causes for years lived with disability with significant effects on quality of life, social isolation, and overall health.^1,2^ Coupled with a forecast of increased hearing loss burden worldwide, the NAS and WHO have urgently recommended that access to hearing evaluation be expanded to meet demand.^1,2^ Compounding this increased demand for hearing services, there is a severe shortage of audiologists globally, particularly in developing countries.^3^ For example, many Southeast Asian and African countries report having less than one audiologist per million population.^3^ In developed countries such as the United States, audiology expertise is also in short supply with recent estimates recommending the supply of audiologists be increased immediately by 50% in order to meet forecasted demand.^4^

Recognizing the gap between supply and demand for hearing evaluations, several novel hearing test delivery solutions have emerged, including hearing screening through the telephone and internet mediums, and portable diagnostic audiometers.^5–10^ However, these assessment methods do not replace the need for comprehensive expert audiologist interpretation of the result. Thus, despite these innovations, we have yet to make significant gains in accessibility for diagnostic hearing function evaluation when audiologist interpretation is limited or unavailable.

Machine learning, a domain of artificial intelligence, has experienced explosive popularity in applications in medicine.^11,12^ The pillars of promise in using machine learning for medical applications are grounded in automating diagnostic and prognostic tasks, as well as deriving novel insights from massive biomedical datasets. Deep learning approaches, a machine learning technique that utilizes multi-layered models called ‘neural networks’ that learn how to independently classify text or images when trained on examples labelled by human experts, have experienced a surge in popularity recently. For example, neural networks have been successfully used to diagnose diabetic retinopathy and skin pathologies with accuracy meeting or exceeding human experts.^13–15^ When used to augment clinical diagnostics and decision making, machine learning techniques may serve as the foundation of innovative solutions for mismatches between supply and demand in health care access.

Given the marked deficit in audiology workforce capacity, an exponential expected increase in incidence and prevalence of hearing loss worldwide, and the success of deep learning in diverse prognostic tasks, a deep learning-based approach for automatic interpretation of audiograms has the potential to be a game-changing innovation. As of the writing of this manuscript, interest in applying machine learning to myriad challenges in hearing loss from auditory to physiology to cochlear implant care delivery has seen exponential growth over the past decade.^16^ With regard to audiometric data specifically, one group has deployed machine learning algorithms to automatically estimate audiogram performance based on online participant testing.^17,18^ However, no prior models have been developed to automatically interpret existing traditional plot-based audiogram data. The purpose of this manuscript is to present a novel deep learning proof-of-concept model that accurately and quickly interprets diagnostic audiograms.

## METHODS

This study was reviewed by the Sunnybrook Health Sciences Center ethics review board and deemed exempt from formal review (Protocol # 044-2019).

**Image Data Acquisition & Augmentation**. 1,007 audiogram reports from adult patients were obtained from the Department of Otolaryngology electronic medical record system at the Sunnybrook Health Sciences Center in Toronto, Ontario, Canada from 2017 to 2019. Audiograms representing normal, conductive, mixed and sensorineural morphologies were included. The audiogram hearing loss morphologies were labelled by expert audiologists from the Department of Otolaryngology as part of routine clinical encounters. In the province of Ontario, practicing audiologists are certified and regulated by the College of Audiologists and Speech-Language Pathologists of Ontario (CASLPO). The audiogram database was a convenience sample with no hearing pathology or diagnosis specifically excluded. The audiogram plots were cropped from the body of the audiogram reports and saved as individual jpeg formatted picture files.

The total audiogram image file database was randomly split 80/20 to segment the image dataset into ‘training’ (n = 806) and ‘hold-out’ validation set (n = 201) subgroups respectively. No images contained within the hold-out set were used in the training set. Well-established image transformation techniques were used to increase the number of images available for training the deep learning model.^19^ Prior to each training epoch (i.e., learning cycles), the algorithm that trains the neural network randomly transforms the image within the bounds of predetermined transformation settings. Transform operations including image rotation, warping, contrast, lighting, and zoom were applied to artificially add natural variation to the audiogram images in our training set (e.g., images taken on an angle). Once trained, the neural network model was used to predict the diagnosis of each image in the hold out set, and those predictions were compared to the true classification to evaluate the accuracy of the trained model.

**Deep Learning Approach**. We tested four neural network models to interpret audiogram plots and learn to predict the classifications that audiologists at our institution made using the same plots.

A neural network model can be trained from scratch, or a previously trained model can be trained (i.e., re-purposed) for a new predictive task, which is known as transfer learning. In general, a model trained using a small training set via transfer learning will perform better than one trained from scratch. We performed transfer learning and compared classification accuracy using four previously trained models (ResNet-32, ResNet-50, ResNet-101, and ResNet-152) with convolutional neural network architectures that were previously trained on the ImageNet (image-net.org) database containing over 14 million images.20–23 The inputs for our model were pre-transformed images of static audiogram plots sized to 500 x 500 pixels. During training, we re-trained every layer because images in the original ImageNet database are dissimilar to audiograms. The model output was a classification of the audiogram image as one of four possible labels: i) normal, ii) conductive loss, iii) sensorineural loss, or iv) mixed loss (**Figure 1**). See the **Online Methods Supplement** for further technical details of the neural network algorithm development.

**Figure 1.**
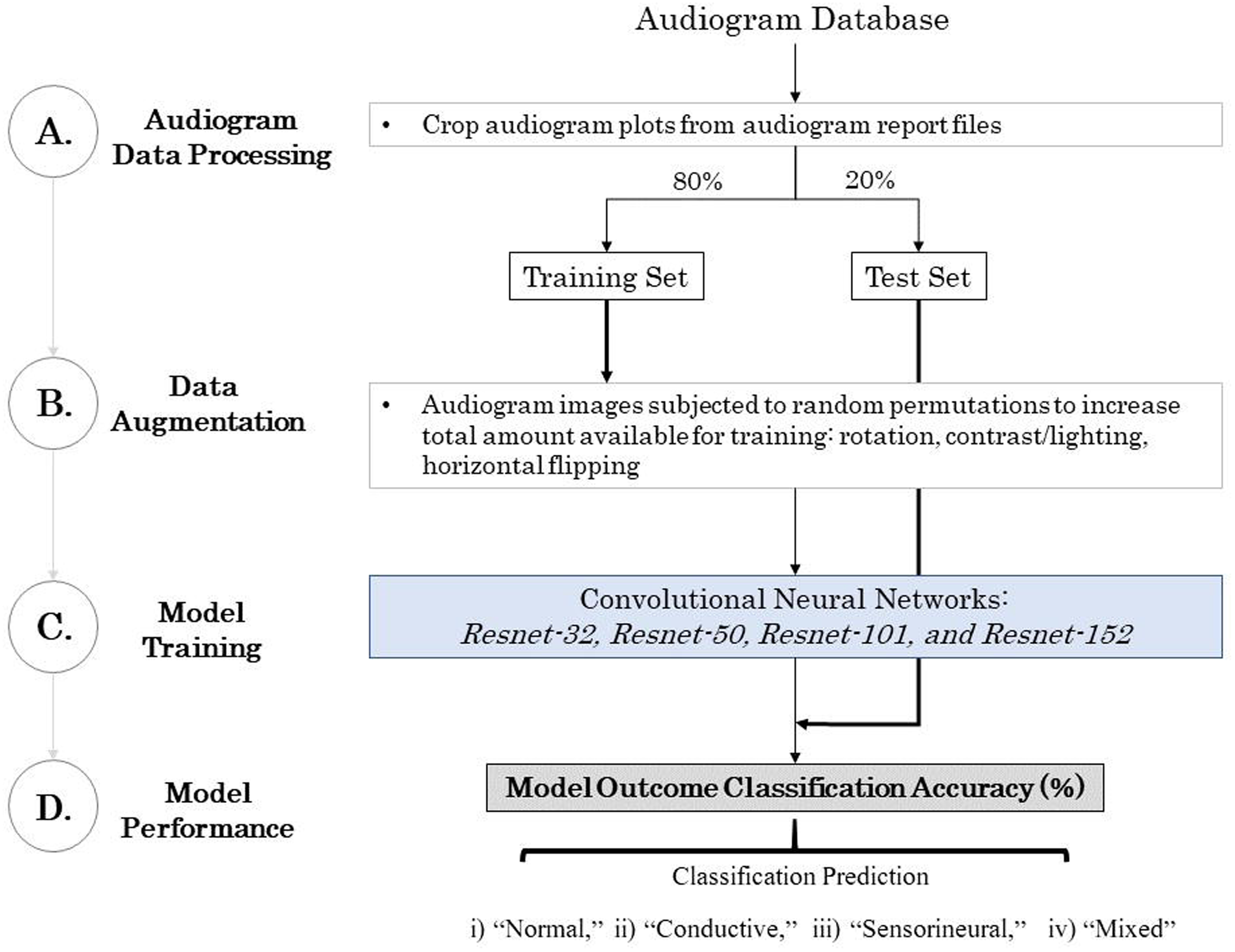
Analytic workflow for preparing and analyzing audiogram plots for automatic morphology classification using neural networks.

**Neural Network Model Prediction Interpretation**. To explore which regions or components of the audiogram images were influential in the neural network arriving at prediction, gradient-weighted class activation heat-maps (Grad-CAM) were generated using the final layer of the trained neural network.^20^ Grad-CAM figures represent a single prediction case with a ‘heat-map’ overlaid on regions of the image that were activated (i.e. largest influence on model output) when predicting the image classification.

## RESULTS

**Model Performance**. The corpus of audiogram images comprised 435 sensorineural, 214 mixed, 207 normal, and 151 conductive hearing loss. The architecture with the highest out-of-training set accuracy was *ResNet-101* at 97.5% (**Table 1**; additional training and performance metrics available in **Online Model Performance Supplemental file**).

**Table 1.** Audiogram interpretation model performance comparison across different neural network architectures based on a hold-out image set.

| **Parameter** | *Resnet-32* | *Resnet-50* | *Resnet-101* | *Resnet-152* |
| --- | --- | --- | --- | --- |
| Layer Depth | 32 | 50 | 101 | 152 |
| Epochs | 30 |  |  |  |
| Learning Rate Range | $1 \times 10^{-7}$ to $1 \times 10^{-2}$ | | | |
| Training Time<br>(minutes:seconds) | 125:24 | 221:30 | 341:12 | 438:23 |
| Model **Test**<br>Classification Accuracy | 96.0% | 97.0% | 97.5% | 97.0% |

Each neural network architecture produced misclassifications that arose from failures of the model to correctly label the audiogram with the appropriate hearing loss type. For *ResNet-32* as an example, the model had the greatest difficulty with true mixed loss morphologies (n = 3 erroneous mixed predictions) and excelled when interpreting normal plots (**Figure 2**). The misclassified audiograms with the highest error rates from *ResNet-50* were extracted and examined using a gradient-weighted class activation heat-map to visualize the model’s ‘interpretation’ of the most important regions of the audiogram for hearing loss morphology (**Figure 3 A-D**). Bright regions in the audiogram plot correspond to regions of the audiogram image where the neural network relied upon for making its label decision. For example, the neural network learned that the separation between the air conduction line and the bone conduction line represented a conductive loss.

**Figure 2.**
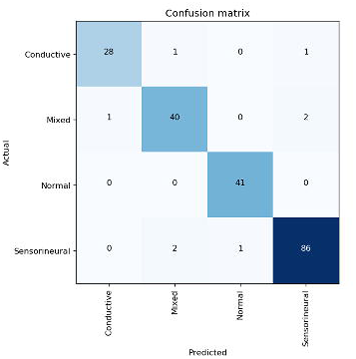
Confusion matrix outlining the class-specific performance of the *ResNet–32* model.

**Figure 3.**
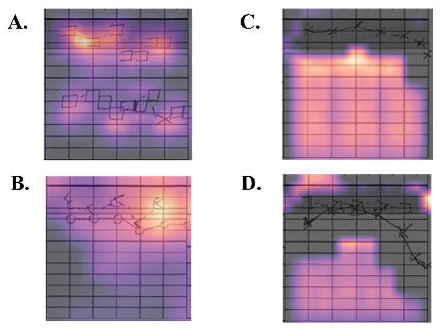
Sample of model misclassifications. **A**. Incorrect classification of a conductive hearing loss, **B**. sensorineural hearing loss, **C**. a normal result, and **D**. a mixed hearing loss.

However, in some instances the model failed to recognize the bone conduction threshold below normal which would classify an audiogram as representing a mixed loss (**Figure 3A**), or a bone conduction threshold value at high frequency that would represent a sensorineural loss, albeit only at one frequency (**Figure 3C**). We emphasize that these predictions were rendered from the morphology of hearing loss data within the audiogram plot and were not informed by any clinical patient factors.

## DISCUSSION

Based on representative audiograms of normal hearing, sensorineural, conductive, and mixed hearing loss, the ‘AutoAudio’ deep learning model was able to autonomously and accurately interpret audiogram plots with 97.5% accuracy. The best performing convolutional neural network architecture was the 101-layer *ResNet* variant, however all *ResNet* variants performed similarly within a two-percent margin.

Similar to how an experienced audiologist interprets audiogram plots, a neural network relies upon spatial relationships inherent to the plot data in order to classify images. Insights into model performance can also be derived from the model failings. The most commonly misclassified hearing loss morphologies were conductive and sensorineural hearing loss when the correct label was mixed hearing loss. Identifying a mixed hearing loss is more conceptually challenging as it requires simultaneous observation of hearing thresholds consistent with both an air and bone conduction loss. Although deeper networks may enhance classification accuracy,^21^ we did not see improved performance in architectures with greater depth. Progressively deeper networks are typically more computationally demanding.^22^ When classification performance is equivocal, a more parsimonious model that requires less training time is preferred.

Classifying a hearing test by morphology alone addresses the fundamental task of determining the type of hearing loss morphology present on audiometric analysis. The diagnosis of hearing loss requires integration of clinical data (e.g. a comprehensive otologic history and physical examination), but a determination of the dominant hearing loss ‘type’ is a key first task. Our model could be deployed to help triage patients with hearing loss for aural rehabilitation candidacy in regions with limited or no audiologist availability after a hearing test is completed using conventional audiometry (**Figure 4a**). Specifically, a local technician could administer the hearing test and the model would instantly provide an interpretation. The model would indicate if further assessment for either amplification or medical evaluation is necessary. A major advantage of our deep learning approach for automatic audiogram interpretation is that this model can work with any existing audiometer software or output, so long as the audiogram plot data is standardized with respect to common frequency and threshold ranges. No new equipment needs to be developed nor is a deviation from specific hearing testing protocol required. This is an important consideration when considering deployment to resource-limited settings. However, we recognize that AutoAudio does not incorporate clinical information nor does it consider other parts of the audiogram test battery such as tympanometry, acoustic reflexes or word recognition scores.

**Figure 4.**
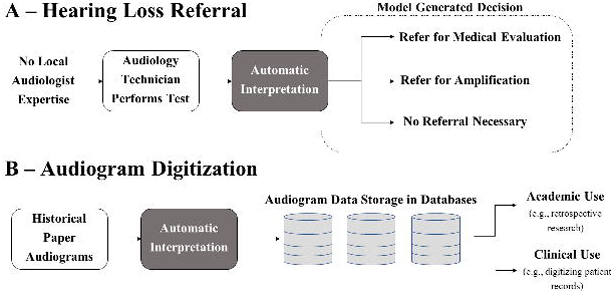
Potential implementations of an automatic audiogram classifier. **A)** In place of a trained audiologist, the automatic audiogram classifier makes a clinical prediction and recommended action. **B)** An automatic audiogram classifier rapidly uploads historic paper audiogram data into a database for use in research or clinical record development.

With the impending increase in hearing loss burden worldwide, there will be a commensurate increase in the need for hearing health services.^2,23,24^ Automatic audiogram interpretation might also confer benefits in developed countries. Audiology stakeholders have recently identified significant bottlenecks and human resource capacity constraints that may limit their ability to meet demand.^4,25^ Re-engineering the hearing evaluation process using machine learning presents an opportunity for generating significant value for audiologists, patients, and health systems. Freed from performing and interpreting routine hearing tests, audiologists may gain time for more pressing tasks such as counselling and aural rehabilitation. Machine learning technology is affordable, scalable, and can be deployed wherever a smartphone or laptop computer can be used. Health systems with adequate audiologist availability may be able to repurpose costs from employing audiologists for hearing evaluations to enhancing or expanding aural rehabilitation programs.

AutoAudio could also be used to swiftly interpret and upload warehoused paper-based audiograms that report hearing performance in the standard plot form (**Figure 4b**). Considering that advances in audiometry and reporting of results may evolve beyond current plot-based result representations, rapid historical audiogram digitization may be AutoAudio’s most durable application. This application may be of interest to research groups looking to rapidly upload old audiogram data at a much faster rate than any human could via manual entry. In a sense, the model is backwards compatible with all audiograms so long as the audiometric data is in plot form. Rapid extraction of audiogram information for inclusion could efficiently build research or clinical databases, or populate a patient’s electronic health record with interpretations of historical non-digitized audiograms.

An alternative approach to automating audiogram interpretation using deep learning is to use rule-based methods such as image understanding through decision trees.^26,27^ In a rule-based method, a human manually defines all features of an audiogram plot deemed predictive of a conductive loss (i.e., presence of an air-bone gap with normal bone conduction thresholds), sensorineural loss (i.e., increased bone conduction thresholds without an air-bone gap), and mixed (i.e. an air-bone gap with increased bone conduction thresholds) hearing loss. One advantage of the rule-based approach is that performance should theoretically approach 100% if the audiogram plot is of sufficient quality for the rules to be evaluated. If audiograms are to be administered in an automated setting in the future with data available in numeric table form - as opposed to a graphical format - this would further facilitate the use of a rules-based algorithm in interpreting audiograms. A major limitation of using numerical frequency values today is that the standard format for recording and storing results on audiometric test performance in many settings is via the audiogram plot. Conversion of audiogram plots to numerical values introduces an additional pre-processing step which would require institutions to change their methodology of reporting for future audiogram plots and more importantly, convert all legacy data to numerical format. Our approach takes advantage of the current reporting standard and could thus be implemented much more easily in practice, since no extra pre-processing is required assuming the audiogram plot format is similar to the plot style the model was trained on.

One of the major disadvantages of a rules-based approach is the requirement for a human to rigorously define the numerical relationships (i.e., air-bone gap, bone conduction thresholds of a mixed loss) of all hearing loss morphologies before being used in the model. Compared to heuristic approaches, a neural network simplifies automatic interpretation as human input is only needed to orient the model to the correct overall classification during the training phase. The neural network ‘learns’ the important features and nuances of audiogram morphologies on its own without any human intervention. One potential weakness in this classification task is that additional model training may be needed for novel images that are significantly different than the image data on which the model was originally trained. However, the model could be easily retrained using heterogenous input images. Furthermore, neural network techniques are becoming widely accessible where limited specialized expertise is needed to train and deploy a model. As was the approach in this study, existing ‘off the shelf’ and open source models can be repurposed for a specific application with relative ease. While it is true that domain expertise and effort could make a comparable performing interpretation model without machine learning, neural networks are easily implemented and powerful. All a hearing test technician would need to implement the neural network model is access to the mobile or computer-based application that can capture the audiogram plot image. There is no need for the additional step of translating the plot data into tabular numerical format needed for a rules-based model, nor to develop new hearing testing equipment or techniques. The neural network is completely backwards compatible so long that the audiogram data is in the standard plot form. In resource-scarce settings, care delivery innovations that do not involve expensive new technologies, additional process steps, or on-site specialized expertise are more likely to succeed.

In the past five years, there have been several notable success stories in using deep learning as a diagnostic tool in cutaneous melanoma and diabetic retinopathy.^13,14^ These models were able to produce classification performance comparable to human ‘experts’ using one neural network architecture - Google’s *Inception V3*.^28^ In this paper, we contribute a novel application to the substantial global health issue of hearing care delivery. To our knowledge, this is the first successful neural network model designed to automate audiogram interpretation. We also demonstrate the model performance gains through testing multiple architectures composed of different functional units and layer depths. In doing so, we were able to generate impressive image classification performance. Our model also takes advantage of enhanced learning rate tuning ‘super convergence’ techniques.^29^ As the field is so new, deep learning model engineering is still a sandbox with no unified path to optimality. Continued experimentation with model constructs and hyperparameters will continue to push model performance to meet and exceed human standards.

There are several limitations associated with our study. First, our model may not be generalizable to audiogram plots that combine both ears on a single plot or use non-standard annotations, scales, and axis lines. Our institution’s audiogram data are hand-written on separate plots for the left and right ears. Future work will focus on training neural network models using audiograms generated from different audiometer software and from collaborating institutions. Second, neural networks generally require large datasets for model training to achieve high accuracy. Our model accuracy would presumably increase with more training data. Third, interpretability in deep learning remains a persistent issue. Feature identification in deep learning models is an active area of research,^30^ and will be key for supporting scaled implementation of such models in clinical applications where confidence in exactly how the models arrive at predictions is needed. Lastly, we did not perform a direct head-to-head comparison between human audiologists and the neural network algorithms. We are planning a follow-up analysis whereby we will challenge AutoAudio against several established audiologists, Otolaryngology residents and audiology trainees. For the purposes of describing the performance of the algorithm in our present paper, an exploration of inter-rater reliability would not affect the performance of the algorithm *per se*. Previously published work has shown that the inter-rater reliability between paired audiologists interpreting air- and bone-thresholds to be within near perfect agreement.^31^ However, exploring inter-reliability assessment on with our local audiologists would be an interesting angle that may support the automation of audiogram interpretation. If significant differences in human audiologist interpretation exist, a case could be made in favor of an algorithm taking over the task from humans to reduce between-operator variability during clinical testing.

Hearing loss is and will continue to be a significant public health issue. Re-engineering the process of hearing testing with a machine learning innovation may help enhance access to the growing worldwide population that is expected to require audiologist services. Our results suggest that deep learning may be a transformative technology that enables automatic and accurate audiogram interpretation. Further work to generate deep learning algorithms that incorporate hearing loss severity, clinical information and the entire audiometric test battery will expand the utility of this approach. Such future models will also need to be trained on audiograms with different graphical formats and annotations to enhance generalizability across different audiometers and practitioners.

## Data Availability

The data that support the findings of this study are available on request from the corresponding author, MGC. The data are not publicly available due to containing information that could compromise the privacy of research participants.

## ACKNOWLEGEMENTS

This work is made possible in part due to the 2018 Connaught Global Challenge Grant Award. The authors would like to thank Jeremy Howard and the team at ‘fast.ai’ for making their technology and educational programming for deep learning inclusive and open to all.

*Specific NIH acknowledgement for D.T.:* the research which is the basis of the paper was conducted prior to *D. T*. starting at the NIH. The content of this paper in no way represents any views held by the NIH.

*M.G.C. designed and tested the model, analyzed data and wrote the paper;*

*T.C. and D.T. provided analytic guidance and critical revision;*

*A.B. and R.M. provided technical oversight and critical revision*.

*A.H., J.L., and V.L. provided critical revision*.

## ONLINE SUPPLEMENT LEGENDS

1. **Online Method Supplement**. Additional methods background on the deep learning model and parameters.
2. **Online Model Performance Supplement**. Additional *ResNet* model performance metrics.

